# The Effect of Incarceration on Tuberculosis Treatment Outcomes in Brazil: a Retrospective Cohort Study

**DOI:** 10.1101/2021.07.31.21261427

**Authors:** Jamieson O’Marr, Crhistinne Gonçalves, Denise Arakaki-Sanchez, Daniele Maria Pelissari, Fernanda Dockhorn Costa, Julio Croda, Katharine S. Walter, Jason R. Andrews

**Author notes:** Correspondence: Jamieson O’Marr, Yale School of Medicine, 333 Cedar St, New Haven CT, USA, 06510. Contributed equally (Julio Croda, Katharine S. Walter, Jason R. Andrews).

## Abstract

**Background:** Tuberculosis notifications in Latin American prisons have more than doubled over the past two decades; however, treatment outcomes and their determinants among incarcerated individuals in this region are not well understood.

**Methods:** Newly diagnosed drug-susceptible tuberculosis cases reported to Brazil’s Sistema de Informação de Agravos de Notificação (SINAN) between January 2015 and December 2017 were included. Multivariate logistic regression was used to assess socioeconomic and clinical factors associated with treatment success among incarcerated individuals.

**Results:** Incarcerated individuals (n=17,776) had greater treatment success than non-incarcerated individuals (n=160,728; 82.2% vs 75.1%, p<0.0001), including after adjusting for demographic and clinical risk factors (adjusted Odds Ratio [aOR]: 1.27; 95% CI: 1.19-1.34). These differences were partially mediated by increased use of directly observed therapy among incarcerated individuals (DOT) (61% vs 47%, p<0.001), which was associated with greater efficacy in the incarcerated population (aOR 2.56 vs aOR 2.17; p<0.001). DOT was associated with improved treatment success among incarcerated subpopulations at elevated risk of poor outcomes.

**Conclusion:** Tuberculosis treatment success among incarcerated individuals in Brazil is higher than non-incarcerated individuals, but both fall below WHO targets. Expanding the use of DOT and services for socially and medically vulnerable individuals may improve outcomes in carceral settings.

## INTRODUCTION

Tuberculosis remains one of the leading causes of death by an infectious disease. In 2019, there were an estimated 10 million cases with approximately 1.2 million deaths among HIV-negative individuals and an additional 250,000 deaths among HIV co-infected individuals^1^. The World Health Organization (WHO) has identified 30 countries with the highest burden of tuberculosis, where 87% of the world’s new tuberculosis cases occurred in 2019^1^. Brazil remains the only country within the Americas within this group.

The incidence of tuberculosis in Brazil has slowly declined over the last 20 years, although in the last three years there has been a modest increase^2^. This trend has been part of a troubling rise in tuberculosis cases across Central and South America in recent years^3^. The rate of incarceration in Central and South America has increased more than 206% since 2000 while tuberculosis cases among incarcerated individuals have increased by an even larger amount^3–7^. Incarcerated individuals represent a disproportionate and growing proportion of the total tuberculosis cases across Central and South America. The higher prevalence of HIV, smoking, alcohol and illicit drug use among incarcerated populations, together with increased risk of infection and deficiencies in health facilities, pose threats to the successful treatment of tuberculosis in correctional settings^4,8,9^.

There are few published studies examining tuberculosis treatment outcomes in correctional facilities in the Americas, and the factors associated with poor outcomes in these settings are not well understood. Understanding these factors well and addressing them will be important for controlling tuberculosis both within incarcerated populations and surrounding communities. We used Brazil’s national tuberculosis registry to measure the effect of incarceration on tuberculosis treatment outcomes and identify risk factors for lack of tuberculosis treatment success among Brazil’s growing incarcerated population.

## METHODS

### Study Design and Participants

We conducted a retrospective cohort study of tuberculosis cases reported to Brazil’s national notifiable disease system, Sistema de Informação de Agravos de Notificação (SINAN), from January 2015 through December 2017. All individuals diagnosed with tuberculosis in Brazil are mandatorily reported to SINAN^10,11^. SINAN reports tuberculosis treatment outcome at the completion of treatment as cure, lost to follow-up, death from tuberculosis, death from other causes, transfer, diagnostic change, drug-resistant tuberculosis, change in treatment regimen, and therapeutic failure.

We excluded patients with a change in diagnosis and those who did not have a reported treatment outcome. Patients who were found to have multidrug-resistant tuberculosis (MDR-TB) cases at diagnosis were also excluded because MDR-TB treatment outcomes are reported in a separate database. MDR-TB cases were defined as those individuals whose *M. tuberculosis* isolate was resistant to isoniazid and rifampin at the time of diagnosis. Additionally, we excluded cases without a reported incarceration status and limited our analysis to only new identified tuberculosis cases due to differing treatment outcome definitions among retreatment tuberculosis cases. Race is a self-reported variable within SINAN with five established survey categories: black, brown or mixed race, white, Asian, and indigenous.

### Outcomes and Definitions

We defined treatment success as cases reported as “cure” in SINAN. The Brazilian National Tuberculosis Program defines cure as two negative sputum smears at the end of therapy or the completion of treatment with no evidence of failure and clinical and radiological criteria in individuals without a sputum smear test. In our models, we defined lack of treatment success as lost to follow-up, death due to tuberculosis, death due to other causes, therapeutic failure (including MDR diagnosis after treatment), or transfer. We included all newly reported tuberculosis cases among people above the age of 18 at diagnosis with complete information for incarceration status and treatment outcome.

In addition to the treatment outcome, the following variables were extracted from SINAN: 1) incarceration status at the time of tuberculosis notification, 2) sex, 3) age at diagnosis, 4) year of diagnosis, 5) self-reported race, 6) education level, 7) received directly observed therapy (at least 3 weekly assisted doses throughout the treatment course), 8) alcohol use disorder, 9) HIV status, 10) diabetes status, 11) presence of a mental health condition, 12) clinical form of tuberculosis 13) Brazilian state of diagnosis. The clinical variables of alcohol use disorder, diabetes, and presence of a mental health condition are determined and reported by the healthcare worker notifying the case to SINAN at the time of diagnosis.

### Statistical Analysis

Data analysis was conducted in R (version 3.5.3) and R Studio (version 1.2.1335). Missingness was under 10% for the majority of the variables included in our analysis and covariates with missingness greater than 30% were excluded. To address missingness in measured covariates, we employed multiple imputation for race, education level, directly observed therapy (DOT), HIV status, alcohol use disorder, diabetes, and the presence of a mental health condition using the mice package in R^12,13^.

We compared our outcomes and extracted demographic and clinical variables between the non-incarcerated and incarcerated population with chi-squared statistics. We tested the effect of clinical and sociodemographic variables on tuberculosis treatment outcome with mixed effects multivariable logistic regression including incarceration status, DOT, sex, self-reported race, age strata, year of diagnosis, education level, HIV status, alcohol use disorder, diabetes, mental health condition, and form of tuberculosis with state included as a random effect. These variables were selected based on previous studies and subject level knowledge of factors that affect tuberculosis treatment outcomes. We included a multiplicative interaction between DOT and incarceration to assess for any relationship between these two variables. Odds ratios with 95% confidence intervals were reported. To investigate the relationship between the predictors in the above model more specifically among the incarcerated population, a multivariable logistic regression was employed.

### Ethics Statement

This study was approved by the IRB at Stanford University (Protocol #50466) and the IRB at the Universidade Federal Do Mato Grosso Do Sul (Protocol #20531819.5.0000.0021).

## RESULTS

A total of 178,504 new tuberculosis cases were reported to Brazil’s national tuberculosis registry between 2015 and 2017. Among those, 17,776 (9.96%) individuals were incarcerated at the time of diagnosis. The overwhelming majority of cases notified in the incarcerated population occurred among men (96.4%), compared to 66.2% of cases notified in the general population (χ^2^ test, p< 0.0001). The proportion of individuals with tuberculosis who were HIV co-infected was lower among the incarcerated population than the general population (6.4% vs 11.1%; p<0.0001), as was the percentage of patients with reported alcohol use disorder (12.7% vs 18.6%, p< 0.001). Incarcerated individuals were younger (30.4 vs 42.7 years, p<0.0001) and were more likely to be treated under DOT than non-incarcerated individuals (61.0% vs 47.4%, p< 0.0001).

135,238 (75.8%) were considered to have treatment success, 18,261 (10.2%) lost to follow-up, 5,714 (3.2%) died of tuberculosis, 8,175 (4.6%) died by other causes, 9,254 (5.2%) transferred from the program, 730 (0.4%) were subsequently diagnosed with drug-resistant tuberculosis after a trial of therapy, 1,012 (0.6%) changed treatment regimen, and 120 (0.07%) experienced therapeutic failure. Treatment success was higher among incarcerated individuals compared with the general population (82.2% vs 75.1%, p<.0001). Notable differences in the remaining treatment outcomes between the incarcerated and non-incarcerated populations was a lower rate of death by tuberculosis (0.9% vs 3.5%; p<0.0001), death by other causes (1.3% vs 4.9%; p<.0001) and lost to follow-up (8.5% vs 10.4%; p<0.0001).

In a multivariable model, incarceration at the time of tuberculosis notification was associated with an increased odds of treatment success (Table 2; aOR: 1.26; 95% CI: 1.19-1.34). DOT was also strongly associated with treatment success (aOR: 2.16; 95% CI: 2.11-2.22). We also found a significant interaction between incarceration and DOT (aOR: 1.18; 95% CI: 1.09-1.28), suggesting that DOT has a greater positive impact on treatment success among the incarcerated population compared to the general population. Factors that were negatively associated with treatment success in the entire cohort included HIV co-infection (aOR: 0.33; 95% CI: 0.32-0.34), reported alcohol use disorder (aOR: 0.55; 95% CI: 0.54-0.57), age greater than 65 years (aOR: 0.71; 95% CI: 0.68-0.75) and self-reported non-white race (black, aOR: 0.80; 95% CI 0.75-0.84; mixed race, aOR: 0.81; 95% CI: 0.79-0.84; Asian, aOR: 0.83; 95% CI: 0.73-0.94).

**Table 1.** Sociodemographic and clinical characteristics of new Brazilian TB patients from 2015-2017 after imputation, by incarceration status.

|  |  | Cohort |  | P value |
| --- | --- | --- | --- | --- |
|  |  | Non-Incarcerated (%)<br>(N = 160728) | Incarcerated (%)<br>(N = 17776) |  |
| Sex |  |  |  | < .001 |
|  | Men | 106516 (66.2) | 17134 (96.4) |  |
|  | Women | 54212 (33.8) | 642 (3.4) |  |
| Mean age (years) |  | 42.7 | 30.4 | < .001 |
|  | 18-25 | 27793 (17.3) | 6809 (38.3) | < .001 |
|  | 26-45 | 66979 (41.7) | 9753 (54.9) |  |
|  | 46-65 | 49552 (30.8) | 1041 (5.8) |  |
|  | > 65 | 16404 (10.2) | 173 (1.0) |  |
| Self-Reported Race |  |  |  | < .001 |
|  | white | 56680 (35.3) | 6250 (35.2) |  |
|  | black | 21341 (13.2) | 2171 (12.2) |  |
|  | Asian | 1297 (0.8) | 164 (0.9) |  |
|  | brown/mixed Race | 79662 (49.6) | 9103 (51.2) |  |
|  | indigenous | 1740 (1.1) | 88 (0.5) |  |
| Education (years) |  |  |  | < .001 |
|  | Less than 5th Grade | 48222 (30.0) | 4439 (25.0) |  |
|  | 5th - 8th Completed | 49177 (30.5) | 8400 (47.3) |  |
|  | Greater than 8th Completed | 63318 (39.5) | 4937 (27.7) |  |
| Directly Observed Therapy |  | 76124 (47.4) | 10844 (61.0) | < .001 |
| Comorbidities |  |  |  |  |
|  | Alcohol Use Disorder | 29836 (18.6) | 2265 (12.7) | < .001 |
|  | HIV Positive | 17918 (11.1) | 1145 (6.4) | < .001 |
|  | Diabetes | 14838 (9.2) | 304 (1.7) | < .001 |
|  | Mental Health Condition | 4141 (2.6) | 276 (1.5) | < .001 |
| Tuberculosis Form |  |  |  | < .001 |
|  | Pulmonary | 132705 (82.6) | 16921 (95.1) |  |
|  | Extrapulmonary | 22834 (14.2) | 667 (3.8) |  |
|  | Extrapulmonary and Pulmonary | 5189 (3.2) | 188 (1.1) |  |
| Treatment Outcome |  |  |  | < .001 |
|  | Cure | 120632 (75.1) | 14606 (82.2) |  |
|  | Lost to Follow-up | 16748 (10.4) | 1513 (8.5) |  |
|  | Death by TB | 5553 (3.5) | 161 (0.9) |  |
|  | Death by Other Cause | 7946 (4.9) | 229 (1.3) |  |
|  | Transfer | 8045 (5.0) | 1169 (6.6) |  |
|  | MDR-TB | 661 (0.4) | 69 (0.4) |  |
|  | Change in Treatment regimen | 986 (0.6) | 26 (0.1) |  |
|  | Therapeutic Failure | 117 (0.1) | 3 (0.02) |  |

**Table 2.**
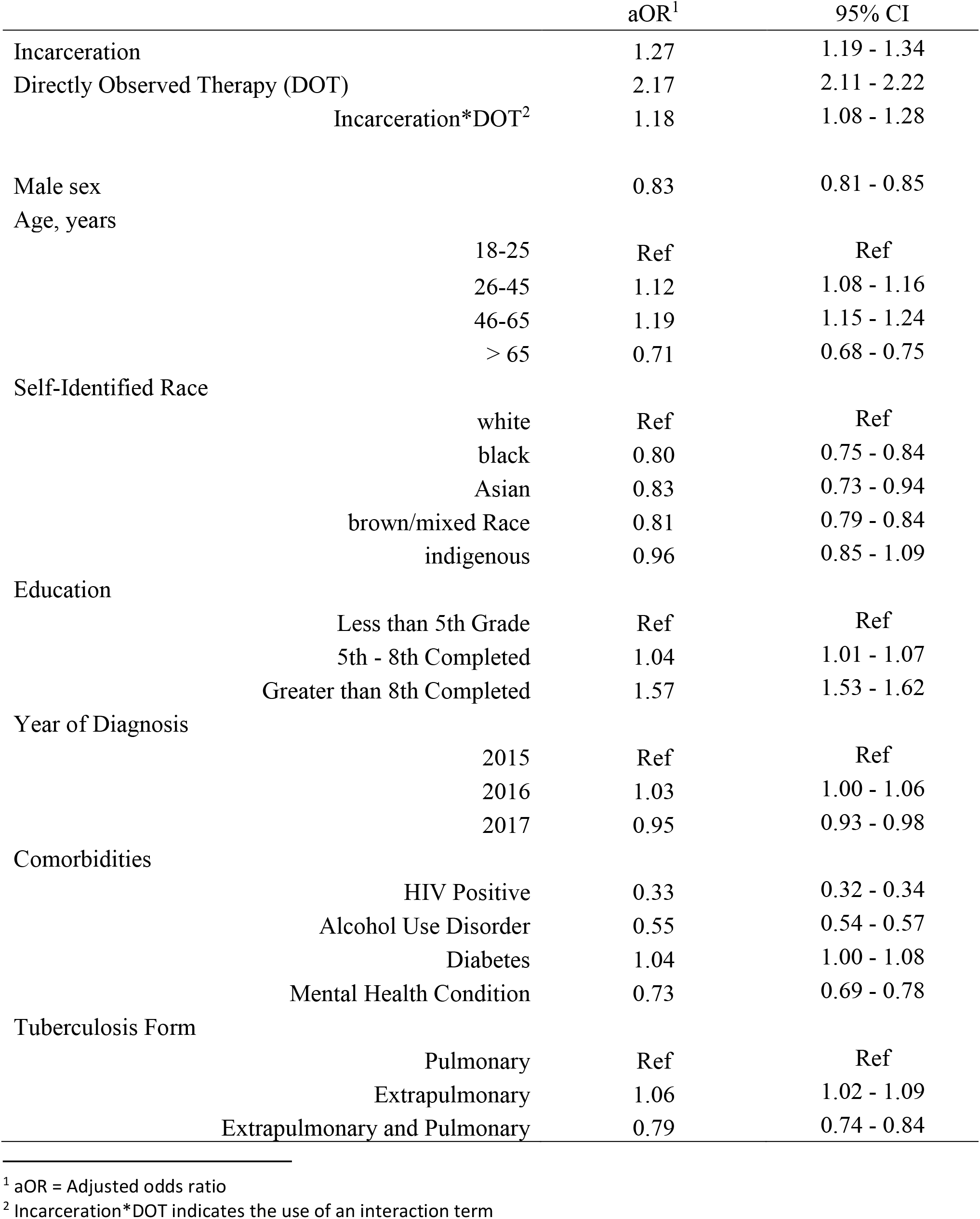
Adjusted odds ratios for treatment success among all newly diagnosed tuberculosis cases using mixed-effects multivariable regression.

|  | aOR<sup>1</sup> | 95% CI |
| --- | --- | --- |
| Incarceration | 1.27 | 1.19 - 1.34 |
| Directly Observed Therapy (DOT) | 2.17 | 2.11 - 2.22 |
| Incarceration*DOT<sup>2</sup> | 1.18 | 1.08 - 1.28 |
| Male sex | 0.83 | 0.81 - 0.85 |
| Age, years |  |  |
| 18-25 | Ref | Ref |
| 26-45 | 1.12 | 1.08 - 1.16 |
| 46-65 | 1.19 | 1.15 - 1.24 |
| > 65 | 0.71 | 0.68 - 0.75 |
| Self-Identified Race |  |  |
| white | Ref | Ref |
| black | 0.80 | 0.75 - 0.84 |
| Asian | 0.83 | 0.73 - 0.94 |
| brown/mixed Race | 0.81 | 0.79 - 0.84 |
| indigenous | 0.96 | 0.85 - 1.09 |
| Education |  |  |
| Less than 5th Grade | Ref | Ref |
| 5th - 8th Completed | 1.04 | 1.01 - 1.07 |
| Greater than 8th Completed | 1.57 | 1.53 - 1.62 |
| Year of Diagnosis |  |  |
| 2015 | Ref | Ref |
| 2016 | 1.03 | 1.00 - 1.06 |
| 2017 | 0.95 | 0.93 - 0.98 |
| Comorbidities |  |  |
| HIV Positive | 0.33 | 0.32 - 0.34 |
| Alcohol Use Disorder | 0.55 | 0.54 - 0.57 |
| Diabetes | 1.04 | 1.00 - 1.08 |
| Mental Health Condition | 0.73 | 0.69 - 0.78 |
| Tuberculosis Form |  |  |
| Pulmonary | Ref | Ref |
| Extrapulmonary | 1.06 | 1.02 - 1.09 |
| Extrapulmonary and Pulmonary | 0.79 | 0.74 - 0.84 |

<sup>1</sup> aOR = Adjusted odds ratio
<sup>2</sup> Incarceration\*DOT indicates the use of an interaction term

In a multivariable model among only incarcerated individuals, DOT (Table 3; aOR: 2.50; 95% CI: 2.31-2.71) and having completed a high school education (aOR: 1.77; 95% CI: 1.58-1.97) were positively associated with treatment success. We additionally identified several factors associated with lower treatment success within this population. These included a non-white race (black, aOR: 0.83; 95% CI: 0.72-0.94; mixed race, aOR: 0.75; 95% CI: 0.69-0.82; indigenous, aOR: 0.49; 95% CI 0.29-0.82), HIV infection (aOR: 0.39; 95% CI: 0.34-0.45), reported alcohol use disorder (aOR: 0.63; 95% CI: 0.57-0.70), a diagnosed mental health condition (aOR: 0.60; 95% CI: 0.46-0.80), and being over the age of 65 (aOR: 0.46; 95% CI: 0.33-0.65).

**Table 3.** Factors associated with treatment success among incarcerated individuals in a multivariable logistic regression model.

|  | aOR<sup>1</sup> | 95% CI |
| --- | --- | --- |
| Directly Observed Therapy | 2.50 | 2.31 - 2.71 |
| Male sex | 1.23 | 1.01-1.49 |
| Age, years |  |  |
| 18-25 | Ref | Ref |
| 26-45 | 1.10 | 1.01 - 1.19 |
| 46-65 | 0.97 | 0.81 - 1.14 |
| > 65 | 0.46 | 0.33 - 0.65 |
| Self-Identified Race |  |  |
| white | Ref | Ref |
| black | 0.83 | 0.72 - 0.94 |
| Asian | 0.69 | 0.45 - 1.05 |
| brown/mixed Race | 0.75 | 0.69 - 0.82 |
| indigenous | 0.49 | 0.29 - 0.82 |
| Education |  |  |
| Less than 5th Grade | Ref | Ref |
| 5th - 8th Completed | 1.31 | 1.20 - 1.44 |
| Greater than 8th Completed | 1.77 | 1.58 - 1.97 |
| Year of Diagnosis |  |  |
| 2015 | Ref | Ref |
| 2016 | 1.06 | 0.96 - 1.18 |
| 2017 | 1.02 | 0.93 - 1.13 |
| Comorbidities |  |  |
| HIV Positive | 0.39 | 0.34 - 0.45 |
| Alcohol Use Disorder | 0.63 | 0.57 - 0.70 |
| Diabetes | 0.97 | 0.72 - 1.30 |
| Mental Health Condition | 0.60 | 0.46 - 0.80 |
| Tuberculosis Form |  |  |
| Pulmonary | Ref | Ref |
| Extrapulmonary | 0.80 | 0.66 - 0.97 |
| Extrapulmonary and Pulmonary | 0.58 | 0.42 - 0.80 |

<sup>1</sup> aOR = adjusted odds ratio

Using predicted probabilities from our multivariable model among incarcerated individuals, we demonstrate that DOT was associated with increased treatment success across a number of important predictors including age, self-reported race, sex, alcohol use disorder, education level, and HIV status (Figure 1). Individuals with risk factors associated with lower probability of treatment success had substantially improved outcomes when treated with DOT, with most subpopulations having an estimated treatment success probability above 85%. Use of DOT among incarcerated populations varied substantially across Brazil, ranging from 0% of incarcerated patients uncovered by DOT in the state of Amapá (n = 47) to as many as 89% of incarcerated patients not utilizing DOT in the state of Rondônia (n = 238) (Figure 2). In total, 8 out of Brazil’s 27 states had over half of their incarcerated population not using DOT, leaving the national average DOT coverage among incarcerated patients at 61%. The states with the largest population of incarcerated individuals not utilizing DOT during tuberculosis treatment were São Paulo (n=2,589), Rio Grande do Sul (n=946), and Rio de Janeiro (n=658).

**Figure 1.**
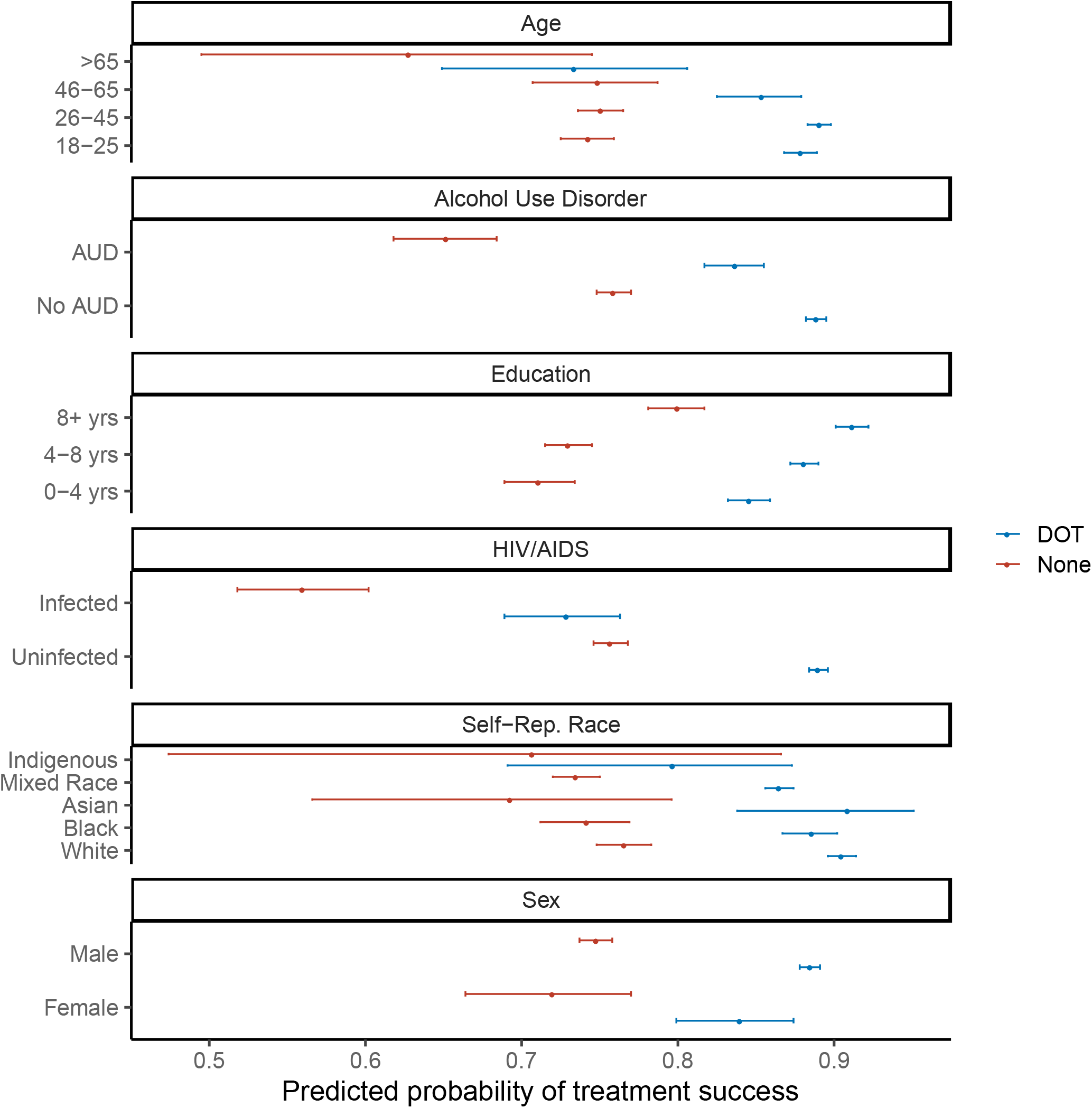
Predicted Probabilities of Treatment Success Among Incarcerated Individuals from the Incarcerated Multivariable Model Stratified by DOT Status. Facets indicate risk factors, points indicate mean predicted probability of treatment success, whiskers indicate interquartile ranges, and color indicates DOT treatment status.

**Figure 2.**
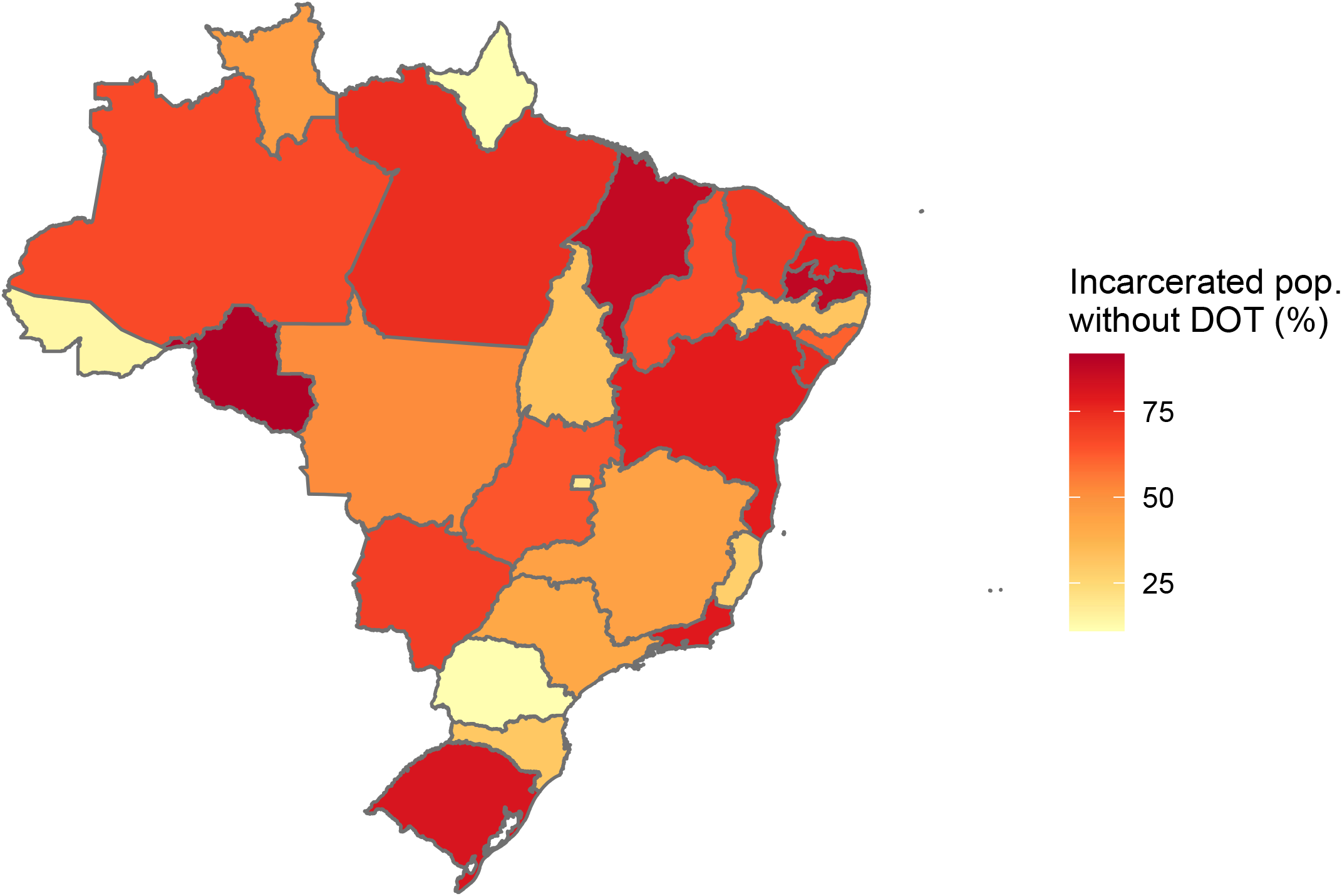
Percentage of the Incarcerated Tuberculosis Patients without DOT by State from 2015 to 2017. Darker colors indicate an increasing percentage of incarcerated tuberculosis patients without DOT.

## DISCUSSION

Tuberculosis notifications have been rising steeply in prisons in Latin America, largely driven by increasing rates of incarceration^3^. However, the impact of incarceration on tuberculosis treatment outcomes and risk factors for treatment failure within prisons have only begun to be identified. Drawing upon Brazil’s national tuberculosis notification system, we found that incarceration at the time of tuberculosis diagnosis was associated with higher rates of treatment success compared with the general population. Use of DOT was greater in prisons than the community and was the strongest predictor of tuberculosis treatment success among incarcerated individuals. However, the proportion of individuals achieving treatment success in prisons (82%) and the general population (75%) do not meet the World Health Organization’s END-TB Strategy 2025 target of 90% treatment success^1^. Among incarcerated individuals, we identified several risk factors and demographic groups in whom treatment outcomes are particularly poor which could inform the development of targeted services and interventions to reduce disparities and improve outcomes.

Our study explored DOT and its relationship to overall treatment success, finding that DOT is strongly associated with better treatment outcomes for all tuberculosis patients. Our analysis also explored the interaction between DOT and incarceration status and demonstrated a significant positive interaction, which underscores the particular importance of DOT for the incarcerated population. Previous literature surrounding DOT has shown mixed efficacy^13^, particularly in correctional settings, however our findings add to the work by Macedo et al. and others which demonstrate a similar beneficial effect of DOT on treatment outcomes among incarcerated individuals in Brazil^14–16^. The usage of DOT within the carceral setting poses unique challenges; however, the observed ability of certain states to employ DOT among nearly all incarcerated tuberculosis patients indicate that it is possible. The skills and strategies for the effective employment of DOT therapy require further study as it has the potential to improve population-level tuberculosis treatment outcomes within Brazilian prisons and in the general population.

Our finding of increased odds of treatment success with incarceration is consistent with previous studies of incarcerated individuals. These studies demonstrate that diagnosing and treating tuberculosis during incarceration, and other infectious diseases such as HIV, may, in some circumstances, improve treatment outcomes due to increased access to medical care during incarceration amongst populations systematically excluded from care^17,22^. However, these studies also find that transitions of care, such as the kinds that result from transfer within a penal system or release, often result in poorer overall outcomes in incarcerated individuals ^17,19–22^. Further studies are needed to investigate outcomes of tuberculosis treatment among individuals who are receiving therapy at the time of release from prison.

Previous studies have identified racial disparities in tuberculosis treatment outcomes in the general population in Brazil.^8,23,25^ We found that such disparities also exist among incarcerated individuals. Adjusting for multiple other demographic and clinical attributes, black incarcerated individuals had nearly 20% lower adjusted odds of treatment success than white incarcerated individuals. Whether this disparity is due to delays in diagnoses, differences in treatment or other factors is not clear from the available data, and further studies are needed to understand the determinants of these disparities so that they can be effectively addressed.

Additional factors that were associated with worse outcomes were alcohol use disorder and HIV positive status, both of which are well-documented predictors for poor treatment outcomes^14^. Finally, other studies have identified the importance of social, educational, and economic support with improved tuberculosis treatment outcomes, something our proxy of education level also supports^17,19^.

While we investigated a large, quality-controlled national tuberculosis registry, our findings have several limitations. First, Brazil’s tuberculosis registry had varying levels of data missingness for several variables. Certain important covariates for treatment success, such as smoking, were missing in more than half of notified cases and had to be excluded. For covariates with less than 30% of observations missing, we used multiple imputation to handle missingness. Another limitation was the lack of standardized screens for clinical covariates such as diabetes, mental health disease, and alcohol use disorder. These determinations were made at the time of diagnosis and thus could be susceptible to bias and underdiagnosis—potentially substantially. Additionally, SINAN lacks economic indicators, limiting us to only education as a social indicator. Due to significant reporting changes in the national tuberculosis registry made in 2015, we could only examine cases from 2015 through 2017. While drug-resistant tuberculosis epidemics in prisons have posed major challenges in many parts of the world, in Brazil, <0.4% of tuberculosis cases in our study were multidrug-resistant which concurs with previous findings by Macedo et al.^14^. Nevertheless, further studies are needed to characterize risk factors for treatment success for multi-drug resistant tuberculosis in correctional settings in Brazil. Despite these limitations, our study utilized a large national surveillance system that enabled a robust evaluation of the covariates associated with treatment success in Brazilian prisons.

There is an urgent need to identify robust strategies for prompt diagnosis and effective treatment to address the increasing burden of tuberculosis in prisons in Brazil and across the Americas. While treatment success rates in prisons exceed those of the general population in Brazil, there remains substantial room for improvement, particularly among vulnerable incarcerated populations. Our findings suggest that tuberculosis treatment success can be improved through expanding the use of DOT, particularly for incarcerated sub-populations at the highest risk of poor outcomes.

## Data Availability

The data used in this article was obtained from from Sistema de Informação de Agravos de Notificação (SINAN) and used at their discretion.

