## Supplemental Table 1 for "The Effect of Incarceration on Tuberculosis Treatment Outcomes in Brazil: a Retrospective Cohort Study"

**Supplementary Table 1**. Sociodemographic and clinical characteristics of new Brazilian TB patients from 2015-2017 before imputation, by incarceration status.

| | Cohort | |
|---|---|---|
| | Non-Incarcerated (%)<br>(N = 160728) | Incarcerated (%)<br>(N = 17776) |
| Sex | | |
| Men | 106511 (66.2) | 17133 (96.4) |
| Women | 54209 (33.8) | 642 (3.4) |
| Missing | 8 (0.0) | 1 (0.0) |
| Mean age (years) | 42.7 | 30.4 |
| 18-25 | 27793 (17.3) | 6809 (38.3) |
| 26-45 | 66977 (41.7) | 9753 (54.9) |
| 46-65 | 49552 (30.8) | 1041 (5.8) |
| > 65 | 16404 (10.2) | 173 (1.0) |
| Missing | 2 (0.0) | 0 (0.0) |
| Self-Reported Race | | |
| Branca/white | 52646 (32.8) | 5612 (31.6) |
| Preta/Black | 19679 (12.2) | 1866 (10.5) |
| Amarela/Yellow | 1136 (0.7) | 131 (0.7) |
| Pardo/Brown | 74344 (46.3) | 8200 (46.1) |
| Indigena/Indigenous | 1597 (1.0) | 58 (0.3) |
| Missing | 11326 (7.0) | 1909 (10.7) |
| Education (years) | | |
| Less than 5th Grade | 35047 (21.8) | 3126 (17.6) |
| 5th - 8th Completed | 36662 (22.8) | 6135 (34.5) |
| Greater than 8th Completed | 48553 (30.2) | 3587 (20.2) |
| Missing | 40466 (25.1) | 4928 (27.7) |
| Directly Observed Therapy | | |
| Yes | 58186 (36.2) | 8782 (49.4) |
| No | 61550 (38.3) | 5354 (30.1) |
| Missing | 40992 (25.5) | 3640 (20.5) |
| Comorbidities | | |

| | | |
|---|---|---|
| Alcohol Use Disorder | | |
| Yes | 28050 (17.5) | 2051 (11.5) |
| No | 124642 (77.5) | 14023 (78.9) |
| Missing | 8036 (5.0) | 1702 (9.6) |
| HIV Status | | |
| Positive | 17000 (10.6) | 1057 (5.9) |
| Negative | 135413 (84.2) | 15364 (86.4) |
| Missing | 8315 (5.2) | 1355 (7.6) |
| Diabetes | | |
| Yes | 14033 (8.7) | 268 (1.5) |
| No | 138334 (86.1) | 15754 (88.6) |
| Missing | 8361 (5.2) | 1754 (9.9) |
| Mental Health Condition | | |
| Yes | 3835 (2.4) | 237 (1.3) |
| No | 148201 (92.2) | 15797 (88.9) |
| Missing | 8692 (5.4) | 1742 (9.8) |
| Tuberculosis Form | | |
| Pulmonary | 132705 (82.6) | 16921 (95.1) |
| Extrapulmonary | 22834 (14.2) | 667 (3.8) |
| Extrapulmonary and Pulmonary | 5189 (3.2) | 188 (1.1) |
| Missing | 0 (0.0) | 0 (0.0) |
| Treatment Outcome | | |
| Cure | 120632 (75.1) | 14606 (82.2) |
| Lost to Follow-up | 16748 (10.4) | 1513 (8.5) |
| Death by TB | 5553 (3.5) | 161 (0.9) |
| Death by Other Cause | 7946 (4.9) | 229 (1.3) |
| Transfer | 8045 (5.0) | 1169 (6.6) |
| MDR-TB | 661 (0.4) | 69 (0.4) |
| Change in Treatment regimen | 986 (0.6) | 26 (0.1) |
| Therapeutic Failure | 117 (0.1) | 3 (0.02) |
